# Internal/External Bacterial Sources of Subgingival Plaque Reconstitution

**DOI:** 10.1101/2025.01.23.25321046

**Authors:** Andrew Gibb, Anjali Bhagirath, Lavanya Jain, Monica Gibson, Deanna Williamson, Khaled Altabtbaei

**Affiliations:** Division of periodontics. Mike Petryk School of Dentistry. Faculty of Medicine and Dentistry, University of Alberta. Edmonton, Alberta, Canada; Currently in private practice in Edmonton, Alberta, Canada; Department of Periodontology, School of Dentistry. Indianapolis, Indiana, USA; Department of Human Ecology. Faculty of Agricultural, Life and Environmental Sciences. Edmonton, Alberta, Canada; Division of periodontics. Faculty of Dentistry, Dalhousie University. Nova Scotia, Halifax, Canada

**Keywords:** Human Microbiome, Child Development, Dental Prophylaxis, Dental Plaque, Environmental Genomics, Longitudinal Studies

## Abstract

**Objectives:** The objectives were to quantify the contributions of internal (self) and external (familial) sources to the recolonization of the bacterial content of the subgingival plaque following professional prophylaxis and assess the effect of close-contact activities on modifying this contribution.

**Materials and Methods:** Families, each consisting of at least one preschool-aged child and at least one sibling, were recruited for this interventional cohort pilot study. Microbial samples were collected from various oral sites, including saliva, buccal mucosa, tongue, supragingival plaque, and subgingival plaque in all family members. Following the child’s oral prophylaxis, subgingival plaque samples were collected one week later. DNA from these samples was extracted and sequenced using the 16S rRNA gene and estimation of the sources were quantified using Bayesian source tracking models. Additional analyses using generalized linear mixed models, Phylofactorization, and Spearman correlations. Statistical significance was set at p<0.05.

**Results:** Child’s own subgingival plaque was the primary source of recolonization, contributing 63.7% to the microbial community one-week post-prophylaxis. Siblings contributed approximately 8%, a contribution significantly higher than that from parents, who contributed around 3% each (p<0.05). The analysis revealed a statistically significant positive correlation between the number of siblings and their bacterial contribution to the child’s subgingival plaque. Several close contact activities between parents and children were statistically associated with higher contribution (p<0.05, Spearman correlation). Additionally, 110 bacteria were statistically significantly different in their internal contribution compared to external, after accounting for household association, sample type, and family members (p<0.05, Phylofactor)

**Conclusion:** The findings challenge the traditional focus on parent-child transmission of oral microbes, highlighting the importance of studying families as a whole.

## Introduction

Microbial acquisition in children is a critical process that influences their current oral health and their future susceptibility to oral diseases. The establishment and development of the oral microbiome begin early in life, and the surrounding environment and close familial interactions significantly shape its composition (Mason et al. 2018; Monteiro et al. 2015; Monteiro et al. 2021; Monteiro et al. 2014; Reis et al. 2023). The dynamic nature of the oral microbiome means that it is continually influenced by various factors, including genetic predispositions, environmental exposures, and interactions with other individuals, particularly within the family unit.

Recent studies have demonstrated the environment which is most impactful on the acquisition of the oral microbiome. By sequentially studying the similarity in the salivary microbiome from individuals in the same population, to individuals in a family, to mother-and-child dyads, Valles-Colomer et al. demonstrated that familial membership is the most consequential environment in salivary microbial similarity (Valles-Colomer et al. 2023). Interestingly, the study also included a dataset of individuals living in relatively isolated populations in the Fiji Islands, thereby allowing investigation of microbial similarities not obfuscated by frequent human contact with unfamiliar individuals(Brito et al. 2019; Valles-Colomer et al. 2023). Cohabiting individuals showed a median oral strain-sharing rate of 32%, significantly higher than the 3% in non-cohabitating individuals in the same population and the 0% in non-cohabitating individuals in different populations. Similar microbiota were found in mother-child pairs and within spousal pairs, suggesting that the largest contributor is person-to-person close-contact transmission or a shared environmental source(Valles-Colomer et al. 2023). This is especially pertinent as these spouses were not genetically related. Notably, the duration of cohabitation was positively associated with strain sharing, with a more robust statistical signal than the age or genetic relatedness of the individuals (Valles-Colomer et al. 2023). This close contact-mediated microbial sharing pattern was evident even when this close contact only started in adulthood (e.g., 38% of oral strain sharing between partners). Interestingly, this strain sharing was partially reversible over time; twins who lived apart for 30 years showed a decrease in strain sharing from 30% to about 10% (Valles-Colomer et al. 2023). The same patterns were seen when compared to people in the United States (Valles-Colomer et al. 2023). Gram-negative species were most highly transmitted in households, compared to mother-child transmission and intra-population sharing (Valles-Colomer et al. 2023). This illustrates that families are the most critical environment for salivary microbial transmission.

In contrast, the Mars simulation study, which placed six individuals in an isolated environment for an extended period, showed that while the oral microbiome of each individual remained relatively stable over time, it was also subject to dynamic changes driven by environmental factors(Bacci et al. 2021). The study found that although the overall diversity of the microbiome decreased in isolation, the remaining microbiota exhibited individual-specific dynamics, suggesting that while the environment shapes the oral microbiome, individual factors also play a significant role.

Previous research on microbial transmission has also shown that the transmission of specific oral microbes is not uniform(Lamell et al. 2000; Preus et al. 1994; Tuite-McDonnell et al. 1997). For instance, the bacteria *Aggregatibacter actinomycetemcomitans* (A.a.) is more commonly transmitted between children and their parents, while *Porphyromonas gingivalis* (P.g.) is more likely to be shared between spouses(Asikainen and Chen 1999; Asikainen et al. 1987; Van Winkelhoff and Boutaga 2005). The rates of transmission for these bacteria vary significantly, with vertical transmission (from parent to child) being more common for A.a. and horizontal transmission (between spouses) being more prevalent for P.g(Van Winkelhoff and Boutaga 2005). This uneven pattern of transmission indicates that microbial transmission is influenced by both the specific characteristics of the bacteria and the nature of the relationships between individuals.

Despite the growing body of knowledge about microbial transmission, significant gaps remain regarding the transmission dynamics of the oral microbiome within families. Specifically, there is limited data on the contribution of non-parent household members, such as siblings, to the subgingival microbiome of children. To our knowledge, this study is the first to quantify the contributions to subgingival plaque from internal sources, such as the child’s oral bacteriome, and external sources, including other family members. By analyzing the contributions of different family members, including siblings, to the recolonization of the subgingival bacteriome, this research provides a more comprehensive understanding of how the familial environment influences oral microbial acquisition. As well, the study identifies specific factors, such as close physical contact and the sharing of oral niches, that contribute to the transmission of oral microbes within the family unit.

## Materials and methods

### Study Design and Participants

The study was approved by the University of Alberta Research Ethics Board (Project No. Pro00120887). Study conforms to STROBE protocol. This 1-week interventional cohort pilot study recruited 13 families, each containing at least one preschool-aged child (five years old or younger) and at least one sibling. We chose this age for the youngest child (termed “test participant” from hereon) to reduce outside-world contact associated with attending schools. Families were recruited from local community organizations such as churches and parent groups. **Sample Collection** During the initial visit to the Oral Health Clinic at the University of Alberta, samples were collected from various oral sites of all family members. Participants were advised not to eat/drink for at least 30 minutes prior to the appointment. The following procedures were used for sample collection:

- <u>Saliva:</u> Unstimulated saliva samples were collected from adults using sterile tubes. The Micro·SAL™ device (Oasis Diagnostics, Vancouver, Washington, USA) was used for children. The device was placed in the mouth for a set period to absorb saliva when the sampler was saturated, after which saliva was extruded from the cotton and stored at −20°C.
- <u>Buccal Mucosa and tongue:</u> The inside of the cheek (buccal mucosa) was sampled using the DNA•SAL™ device (Oasis Diagnostics). Participants were asked to scrub the buccal mucosa and store it in the solution provided with the device at −20C. Cells were removed from the device by vortexing in 1X PBS and lysozyme for 1 hour. The supernatant was then used for downstream DNA isolation. Tongue samples were collected similarly.
- <u>Supragingival Plaque</u>: Supragingival plaque was collected using sterile paper points, which were gently placed at multiple supragingival sites and held in place for 20 seconds. All collected points were pooled in a single tube containing RNAlater for storage at −20°C.
- <u>Subgingival Plaque</u>: Subgingival plaque was collected from the distal surfaces of the lower central incisors using sterile paper points. The points were inserted subgingivally into the sulcus for 20 seconds, then transferred to a tube containing RNAlater for preservation and storage at −20°C.

Following the baseline sample collection, each family’s test subject underwent professional oral prophylaxis to disrupt the existing biofilm in the oral cavity. This procedure involved a comprehensive cleaning of the teeth and gums using rubber cups and pumice to disrupt and remove the existing biofilm, effectively displacing the oral bacteriome. A questionnaire (supplementary) was given to the parents to document familial demographic characteristics, and close contact activities among the family members. Subgingival samples were collected from the test participant one week after the prophylaxis using the same procedure described above.

### DNA Extraction and Sequencing

DNA was extracted from the collected samples using the QIAGEN QIAamp DNA Mini Kit (QIAGEN, Germantown, MD, USA), following the manufacturer’s protocol. The concentration and purity of the extracted DNA were assessed using a Qubit 4 Fluorometer (ThermoFisher Scientific, Waltham, MA, USA) with the Qubit™ 1x dsDNA High-Sensitivity Assay Kit, ensuring that the DNA was of sufficient quality for subsequent sequencing. The V1-V3 regions of the 16S rRNA gene were amplified using specific primers (27F: 5’-ACACTCTTTCCCTACACGACGCTCTTCCGATCTGAAKRGTTYGATYNTGGCTCAG-3’ and 519R: 5’-GTGACTGGAGTTCAGACGTGTGCTCTTCCGATCTACGTNTBACCGCDGCTGCTG-3’). The amplified DNA was then sequenced at the Genome Quebec core facility using high-throughput sequencing technology. One of the one-week subgingival samples did not amplify and therefore was not sequenced. Therefore, the findings in in this manuscript are based on data from 12 families.

### Data Processing and Analysis

The raw sequence data were processed using FAVABEAN, an open source developed by the corresponding author’s lab (https://github.com/khalidtab/favabean). It integrates primer trimming (cutadapt(Martin 2011)), DADA2 parameter identification (Figaro(Weinstein et al. 2019)) and ASV generation (DADA2(Callahan et al. 2016)) into a fully automated pipeline. The DADA2 steps integrated into the pipeline are as follows:

#### 1. Quality Filtering

Low-quality reads were filtered out to ensure that only high-quality sequences were retained for further analysis.

#### 2. Denoising

The DADA2 algorithm was employed to model and correct errors in the sequencing data, resulting in the generation of exact amplicon sequence variants (ASVs). This step involved constructing an error model specific to the data and applying it to remove errors from the sequences.

#### 3. Chimera Removal

Chimeric sequences resulting from the erroneous joining of two unrelated DNA fragments were identified and removed from the dataset.

#### 4. ASV Generation

The final set of ASVs was generated and were used as data for the source tracking.

#### 5. Taxonomy assignment

taxonomy assignment was done against eHOMD V15.22(Chen et al. 2010) via DADA2 naïve Bayesian assignment method where sequences are assigned a taxonomy based on their lowest common ancestor. Species assignment was done on only ASV sequences that matched 100% of the reference database. This was used as input for Phylofactorization(Washburne et al. 2019) to examine if there is a taxonomic rank-based relationship to the transmission.

### Bayesian Source Tracking

To determine the contributions of different family members to the recolonization of the child’s subgingival plaque, Bayesian source tracking model (SourceTracker2(Knights et al. 2011)) was used in the default settings using ASVs. Three different models were used:

- **<u>Model 1</u>**: Contribution from “Unknown” sources by running the algorithm with only test participant’s subgingival samples. The one-week subgingival sample served as “sink”, and their baseline samples were set as “sources”.
- **<u>Model 2</u>**: The one-week test participant’s subgingival sample was assigned as a “sink;” all other baseline samples from the test participant and their family members were set as “sources.”
- **<u>Model 3</u>**: Supplementary Figure 3 shows how each participant in the study had their saliva set as “sink” and all their other samples as “sources.” Only the samples from the first visit were used for this analysis.

### Statistical Analysis

The significance of the contributions from different family members and oral sites was assessed using Spearman correlation and linear mixed-effects models done in R. The mixed-effects models used random effects on a family membership and sample type, with the sample source being a fixed-effect variable.

## Results

<u>Patient Characteristics</u> A summary of the demographic and clinical characteristics of the participants is outlined in Table 1. All test participants were in the primary or mixed dentition phase.

### Sources of Re-establishment of the Subgingival Bacteriome

Alpha rarefaction curves (QIIME2(Bokulich et al. 2018)) of ASVs show adequate coverage of the samples (supplemental figure 1). First, we wanted to know if adding other non-test participant samples contributed to additional information of source sampling that would not be accounted for otherwise. Data are reported as the median[25^th^-75^th^ percentile]. When only the test participant’s sources were used, unknown sources contributed 11.97%[10.23-15.66%], while unknown sources contributed 6%[4.49-8.12%] when all familial samples were included in the modelling, This difference was statistically significant (p-value= 0.0004883, Wilcoxon signed-rank exact test). Therefore, all familial samples were used in the modeling for the rest of the analyses.

One week post-prophylaxis, the primary source of the recolonized subgingival bacteriome was the test participant’s own bacteriome (63.7%[60.4725-75.39%]). The pre-existing subgingival plaque contributed a median of 53.11%[42.94-66.50%] to the newly established bacterial community (Figure 1). This indicates that despite the “ecological catastrophe” induced by the prophylaxis, the subgingival plaque retains a substantial reservoir of microbes that can recolonize the niche. The next most significant contributor was the pre-existing supragingival plaque from the same test participant, which accounted for 8.34%[4.705-17.58%] of the recolonized bacteriome, followed by the buccal mucosa at 0.91%[0.355-1.205%]. These findings suggest a hierarchical recolonization process where the subgingival and supragingival niches play leading roles, with minor contributions from saliva and tongue (Supplementary Figures 2, and 3 for further analysis and evidence).

**Figure 1:**
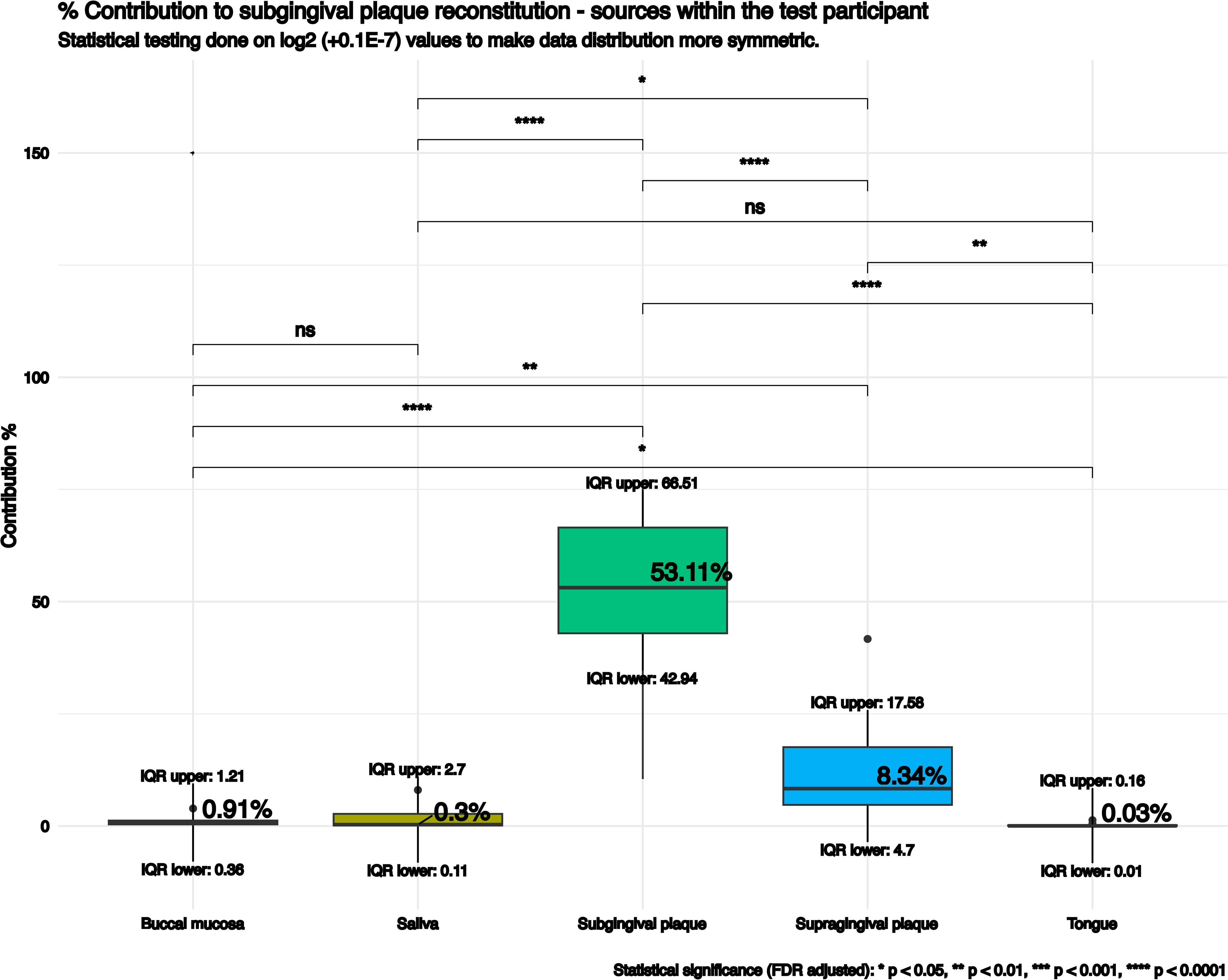
Total contribution of intraoral sources of the child to the subgingival microbiome of the child.

**Figure 2:**
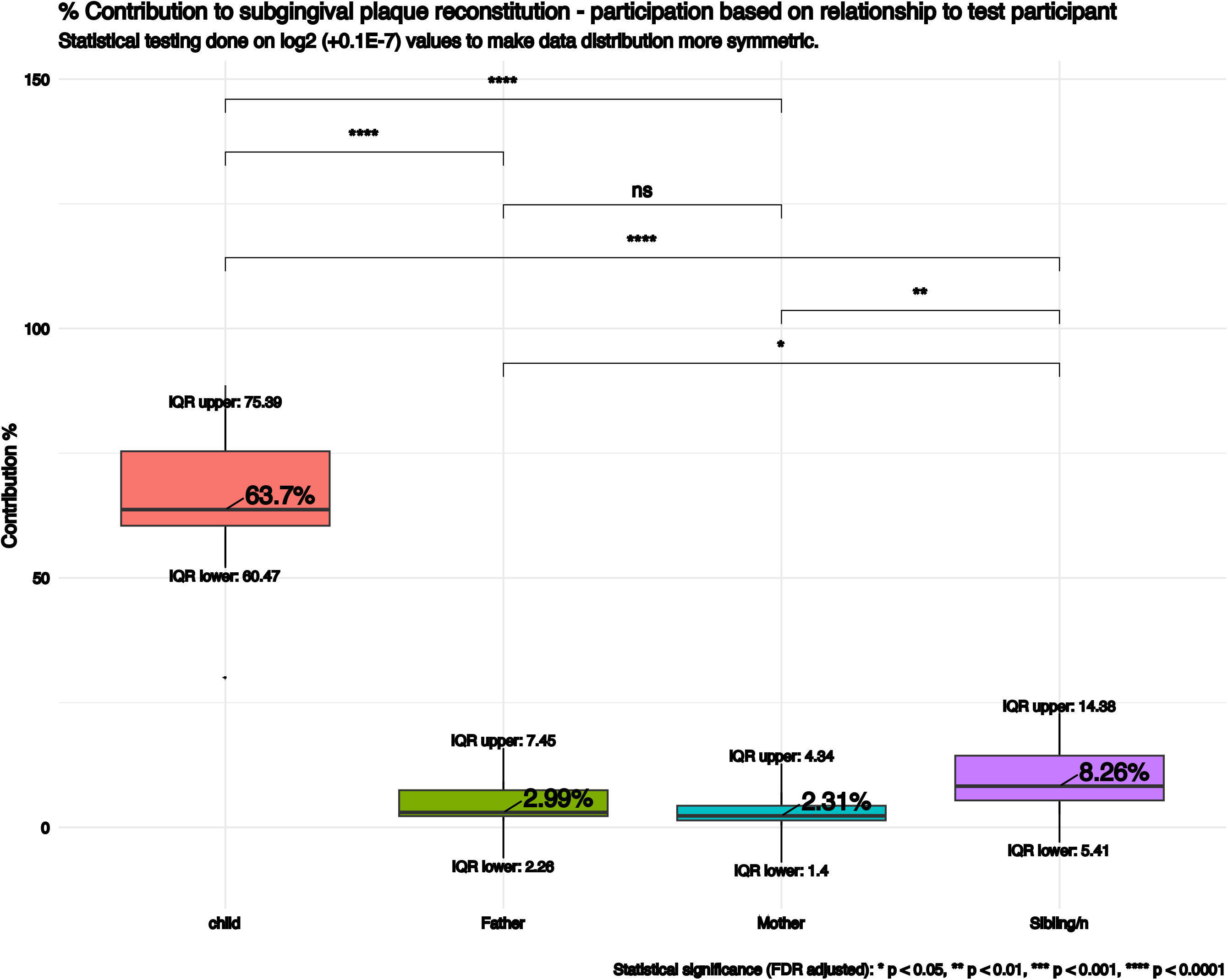
Total contribution to the subgingival plaque of the child based on their relationship to the child.

Interestingly, non-test participant sources also contributed to the recolonization of 27.72%[18.57-32.92%](Figure 3). To account for differing sibling numbers in each family, we divided the contribution by the number of siblings to achieve a “standardized sibling” value (Sibling/n). Sibling/n contributed approximately 8.26%[4.41-14.38%], a figure significantly higher than the contributions from both parents, who each contributed slightly less than 3%. Given these unexpected results, further investigation was done (supplementary table 1). There was a statistically significant positive association between the number of siblings and the bacterial contribution to the test participant’s subgingival bacteriome (Linear mixed model, Estimate = 0.5094, Standard Error = 0.1381, t-value = 3.688, p = 0.000493). Therefore, the larger the number of siblings, the more their collective contribution to the test participant’s subgingival bacteriome, further emphasizing sibling interactions’ role in shaping young children’s oral bacteriome.

**Figure 3:**
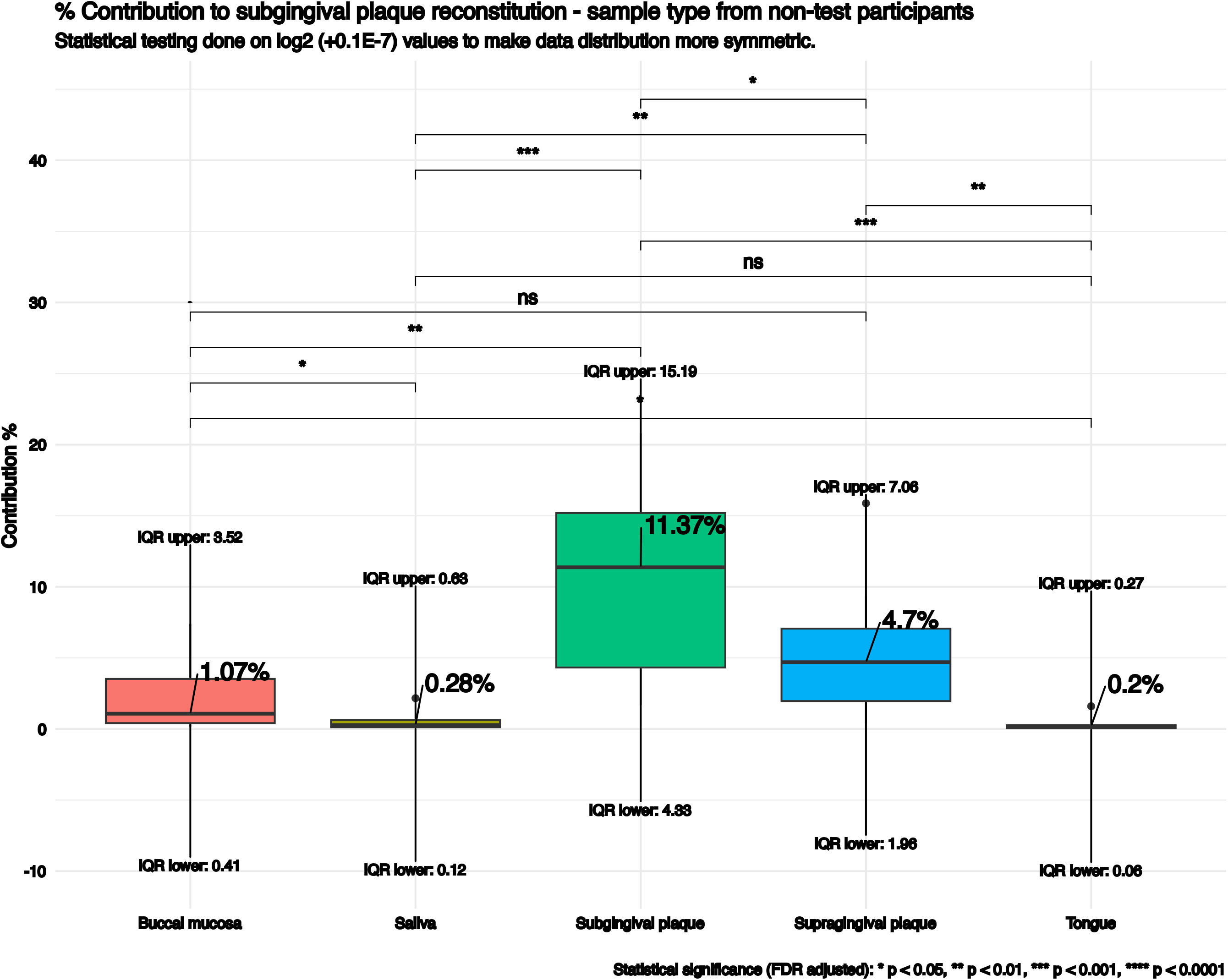
Total contribution to the subgingival microbiome of the child by sample type from all family members.

### Close Contact Activities and Bacterial Transmission

Questionnaire findings showed very few statistically significant associations between close contact activities and parental bacterial contribution, despite the fact that most families (64%) reported frequent close contact activities, such as co-sleeping and shared meals. However, two significant correlations between the reported close-contact activities and the contribution of family members to the test participant’s subgingival bacteriome were noted. The mother’s habit of close sleeping in the vicinity of the test participant was associated with higher mother salivary contribution to subgingival plaque compared to other family member’s salivary transmissions (Rho = 0.641, p = 0.0247). Similarly, the father’s practice of kissing the test participant on the cheek was correlated with the transmission of buccal mucosal microbes to the subgingival plaque (Rho = 0.714, p = 0.009).

### Transmission of Taxa is Not Uniform

Phylofactorization identified 110 statistically significant points on the phylogenetic tree where the contribution of non-test participant’s sources could be observed (Figure 4). Six taxa were predominantly sourced from non-test participant’s sources, even after controlling for the effects of family membership and sample type. These taxa included *Streptococcus mitis, Leptotrichia* sp. HMT_392, *Actinomyces* genus, *Prevotella* genus, *Ruminococcaceae G1* genus, and *Ruminococcaceae* family. More importantly. family membership significantly influenced 92 taxonomic ranks (p<0.05, ANOVA-II). The taxa affected by family membership included several genera such as *Streptococcus, Tannerella, Prevotella*, and *Fusobacterium*. In certain families, the bacterial contribution from siblings was statistically fully aliased by their family membership, indicating a strong familial signature in the bacterial communities of children.

**Figure 4:**
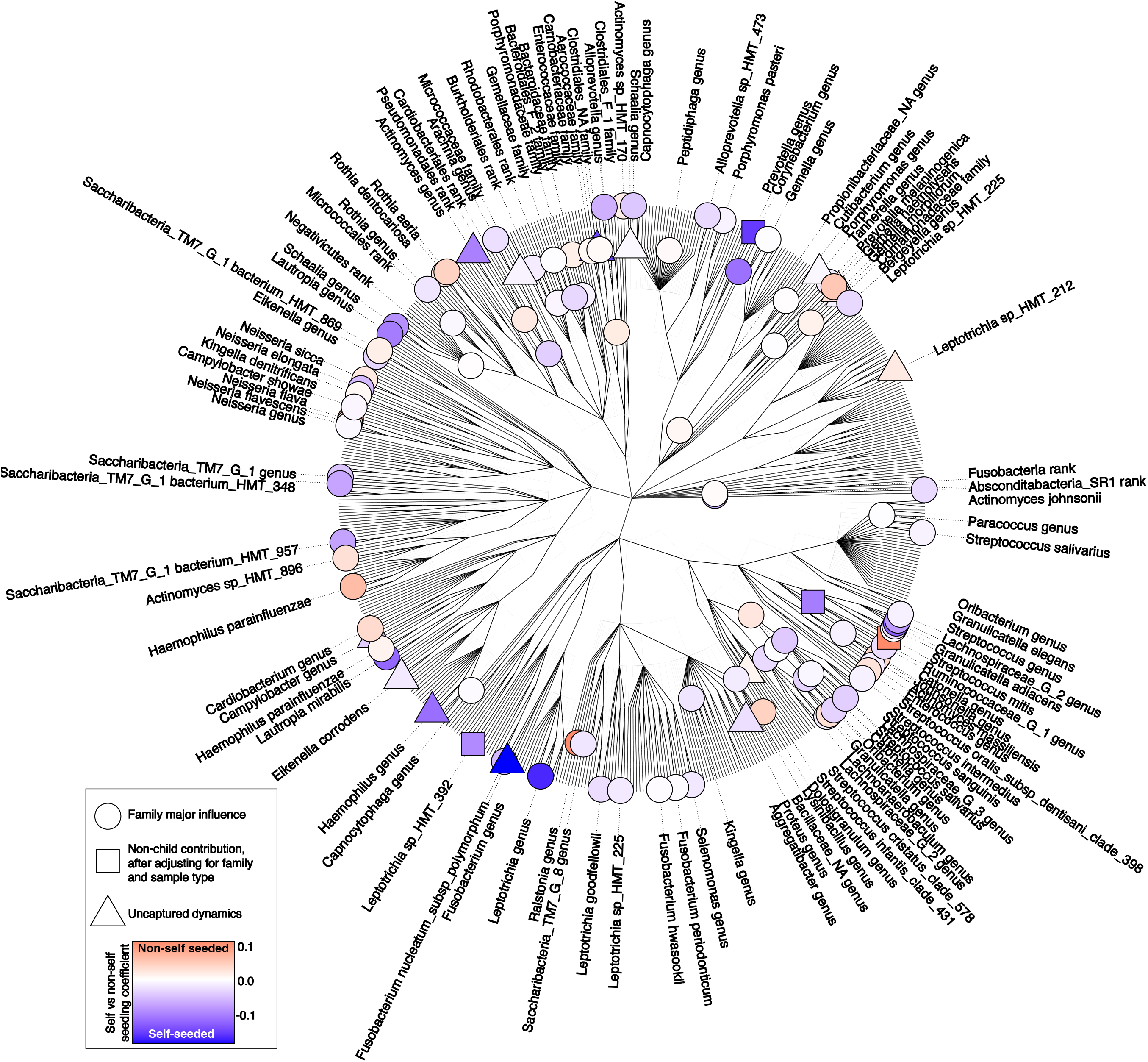
Phylofactorization analysis showing statistically significant points on the phylogenetic tree where family membership influenced bacterial taxa.

## Discussion

This study provides a comprehensive exploration of the bacterial dynamics within the family unit, particularly focusing on the recolonization of the subgingival microbiome in children following professional oral prophylaxis. To our knowledge, it is the first prospective study to estimate the internal/external contributions to the subgingival plaque using Amplicon Sequence Variants (ASVs). Moreover, we believe it is the first prospective study to consider the contributions of siblings to the reconstitution of the microbiome.

External seeding played a significant role in the newly created subgingival microbiota, at 27.72%[18.57-32.92%]. Moreover, findings indicate that siblings, rather than just parents, play a role in shaping a child’s subgingival bacteriome. These findings challenge the traditional view of oral microbiome transmission being vertical (from parents to children). In our study, horizontal transmission (from siblings) plays a larger role than vertical transmission. This may not be surprising as many of the participating families provided care to their youngest child at home, and the study was conducted during the COVID-19 pandemic during a time where children vaccinations were not available which likely meant that siblings were in the household for extended periods of time, be it due to having their education through virtual classrooms, or else. We did not capture this information specifically, especially with respect whether the education was provided in close proximity. However, based on the timeline of return-to-in-person education and reopening in the province, we were able to infer that at least extended periods of time were spent within the household compared to non-COVID-19 periods. Therefore, the children potentially have had a substantial amount of time together. Furthermore, familial membership played a significant role on the transmission at a taxa level. This is not surprising as the collective composition of the bacteriome differs from one family to another; therefore, for a specific taxon to show significance, it must be consistently transmissible across most family structures. These findings suggest that the family membership, with its shared behaviours, genetics and family-specific bacterial profiles, plays a critical role in shaping the test participant’s subgingival bacteriome. This study illustrates two crucial ecological principles: 1) Niche selection and 2) Invasion enhanced by dispersion events. Our study has shown that subgingival bacterial acquisition follows the ecological principle of selection; that is, niche similarity, not phylogeny nor geography, governs the acquisition of subgingival microbiota. This phenomenon has previously been seen in horizontal gene exchanges in human-related microbiomes across various interconnected networks with different reference frames, such as geographic location, individuals, and body sites. Body sites had the most extensive genetic exchange, likely due to their similar niches (niche selection) (Smillie et al. 2011). Our study corroborates that sample types (niches) are the main driver of microbial differentiation between samples (supplementary figure 2), and not family membership and relatedness to test participants. It can be assumed that foreign subgingival plaque is more readily implantable into a newly created subgingival niche due to the similarity in their microbial content.

The second ecological principle illustrated by this study is dispersion, one of the four recognized events that enhance microbial invasion(Kinnunen et al. 2016). When an environment has low abundance, it is more amenable to foreign microbial invasion. A low-abundance environment could occur if the resident microbiota recently underwent a major dispersal event(Kinnunen et al. 2016). Indeed, the purpose of mechanical subgingival plaque disruption is to convert the already established biofilm to one with a low abundance of species in the hope that the biofilm will recreate itself into a more health-compatible biofilm. Therefore, this study proposes that subgingival plaque disruption should be seen as a significant microbial event guided through environmental control rather than undergoing unguided reconstitution. One obvious target is controlling intimate, routine interactions between individuals within the family.

We specifically recruited children at a primary/mixed dentition stage, a consequential period for oral microbial development, in which many of the microbial constituents at this stage can also be seen later in life(Mason et al. 2018; Monteiro et al. 2021). While we followed the participants for only one week, it is possible that further maturation of the biofilm would reduce the external contributions as the resident microbiota regain control over their niches. We suspect this is only partially true as dispersion events occur daily through tooth brushing, which has been shown to enhance the microbial similarity between twins(Freire et al. 2020). Therefore, we believe regular oral hygiene likely increases the external acquisition process through repeated dispersal events, further strengthening the argument that familial close contact should be investigated further. Other possible consequential events such as antibiotics usage by any of the family members prior or during the study, or dietary habits/changes have not been tracked or standardized. Moreover, the relatively homogenous family structures, and the short period of investigation limits the generalizability of this study.

In conclusion, this study advances understanding of the transmission dynamics of the oral bacteriome within families, particularly emphasizing the significant role of siblings. Our findings have important implications: the traditional model of periodontal health and disease as an individual condition may be insufficient to capture the full scope of microbial influences. This study supports a shift toward a family-centered model of investigation, where the oral health of all family members is to be considered. Such an approach could lead to more effective strategies for controlling the spread of pathogenic microbes within families, thereby reducing the overall burden of periodontal disease.

## Supporting information

Contributions

## Data Availability

All data produced in the present study are available upon reasonable request to the authors and will be published in NIH SRA once the publication gets accepted by the journal.

## Acknowledgements

The authors declare no conflict of interest with this manuscript. This work was funded by the Oral Health for Children, Youth, and Families Fund (OHCY-02). We would also like to thank Dr. Mabelle De Freitas Monteiro for her critical appraisal of the analysis, which greatly strengthened this manuscript.

Andrew Gibb – contributed to the conception and design, acquisition, analysis, and interpretation of the data. Drafted the manuscript, and critically revised the manuscript.

Anjali Bhagirath – contributed to acquisition, analysis, and interpretation of the data. Drafted the manuscript, and critically revised the manuscript.

Lavanya Jain – contributed to the acquisition, analysis, and interpretation of the data. Critically revised the manuscript.

Monica Gibson – contributed to the conception and design of the study. Contributed to the interpretation of the data. Critically revised the manuscript.

Deanna Williamson – contributed to the conception and design. Contributed to the acquisition, analysis and interpretation of the data. Drafted the manuscript. Critically revised the manuscript.

Khaled Altabtbaei – contributed to the conception, and design. Contributed to the acquisition, analysis, and interpretation of the data. Drafted the manuscript and critically revised the manuscript.

All authors gave their final approval and agree to be accountable for all aspects of the work.

## Main figure caption list

**Table 1:** Demographic characteristics of the families.

## Notes

### Competing Interest Statement

The authors have declared no competing interest.

### Author Declarations

Ethics committee of The University of Alberta gave ethical approval for this work.

