## Supplementary material for "Internal/External Bacterial Sources of Subgingival Plaque Reconstitution": Contributions

### Statement of Authorship

The submitting author affirms that all individuals listed as authors agree that they have met the criteria of authorship, agree to the conclusions of the study and that no individual meeting the criteria of authorship has been omitted. In order to meet the requirements of authorship, each author must have contributed to at least one aspect of each of the four criteria, as listed below. **Please note that for Criteria 1 and 2, authors only need to meet one of the two items listed**. These criteria are not to be used as a means to disqualify colleagues from authorship who otherwise meet authorship criteria by denying them the opportunity to meet criteria 2 or 3. Therefore, all individuals who meet the first criterion should have the opportunity to participate in the drafting, review, and final approval of the manuscript. Any individuals not meeting the criteria may be mentioned in the Acknowledgements section of the manuscript.

Per the criteria defined by the [International Committee for Medical Journal Editors](http://www.icmje.org/recommendations/browse/roles-and-responsibilities/defining-the-role-of-authors-and-contributors.html) (ICJME), please note the contribution made by each author listed in the manuscript. **Please *type* each role into the boxes that apply. Do NOT respond with “yes or no” or “X” or your form will be returned.**

| Author (Last name, First Initial)  e.g. Smith, J | **Criteria 1** | | **Criteria 2** | | **Criteria 3** | **Criteria 4** |
| --- | --- | --- | --- | --- | --- | --- |
|  | contributed to conception or design | contributed to acquisition, analysis, or interpretation | drafted the manuscript | critically revised the manuscript | gave final approval | Agrees to be accountable for all aspects of work ensuring integrity and accuracy |
| Gibb, Andrew | Contributed to conception and design | Contributed to acquisition, analysis, interpretation | Drafted the manuscript | Critically revised the manuscript | Gave final approval | Agrees to be accountable for all aspects of work ensuring integrity and accuracy |
| Bhagirath, Anjali |  | Contributed to acquisition, analysis, interpretation |  | Critically revised the manuscript | Gave final approval | Agrees to be accountable for all aspects of work ensuring integrity and accuracy |
| Jain, Lavanya |  | Contributed to acquisition, analysis, interpretation |  | Critically revised the manuscript | Gave final approval | Agrees to be accountable for all aspects of work ensuring integrity and accuracy |
| Gibson, Monica | Contributed to conception and design | Contributed to interpretation |  | Critically revised the manuscript | Gave final approval | Agrees to be accountable for all aspects of work ensuring integrity and accuracy |
| Williamson, Deanna | Contributed to conception and design | Contributed to acquisition, analysis, interpretation | Drafted the manuscript | Critically revised the manuscript | Gave final approval | Agrees to be accountable for all aspects of work ensuring integrity and accuracy |
| Altabtbaei, Khaled | Contributed to conception and design | Contributed to acquisition, analysis, interpretation | Drafted the manuscript | Critically revised the manuscript | Gave final approval | Agrees to be accountable for all aspects of work ensuring integrity and accuracy |
