## Supplementary material for "Internal/External Bacterial Sources of Subgingival Plaque Reconstitution": Table 1

| Person | Parameter | | Value | | |
|---|---|---|---|---|---|
| Test participant | Male:Female | Ratio | 5:7 | | |
| | Age (in years) | Mean (STD) | 3.83 (0.94) | | |
| Siblings | # per family | | 3.25 (1.06) | | |
| | Male:Female | Ratio | 3:2 | | |
| | Age (in years) | Count | 5 years | 1 | |
| | | | 6-7 years | 10 | |
| | | | 8-9 years | 6 | |
| | | | 10-11 years | 6 | |
| | | | 12+ years | 2 | |
| **Parents** | | | | **Mother** | **Father** |
| Age (in years) | | Count | 26-30 years | 3 | 2 |
| | | | 31-35 years | 4 | 2 |
| | | | 36-40 years | 5 | 5 |
| | | | 41-45 years | 0 | 2 |
| | | | 46+ years | 0 | 1 |
| Highest educational level obtained | | | <High school | 1 | 0 |
| | | | Highschool | 0 | 1 |
| | | | Some post-secondary | 2 | 1 |
| | | | Diploma (college, trade, technical) | 2 | 2 |
| | | | Undergraduate degree | 0 | 6 |
| | | | Graduate degree | 0 | 2 |
| **Family** | | | | | |
| Familial ethnic affiliation (Note: most reported being mixed ethnicity, therefore; total ethnicity > # of families) | | Count | parents with indigenous ancestry | | 2 |
| | | | children with indigenous ancestry | | 2 |
| | | | Caucasian/European ancestry | | 6 |
| | | | Asian ancestry | | 4 |

1. **Table 1:** Demographic and clinical characteristics of the participants.
